# Dysfunctional Sleep Beliefs and Attitudes in Major Depressive Disorder: Dimensional Associations with Clinical Phenotypes and Gut-Neuroimmune–Metabolic–Oxidative Pathways

**DOI:** 10.64898/2026.10.18.26365736

**Authors:** Min Wang, Mengqi Niu, Xinling Luo, Xiaoli Chen, Hongling Liu, Tangcong Chen, Abbas F. Almulla, Yingqian Zhang, Jia Li, Michael Maes

**Author notes:** Co-corresponding author: Prof. Dr. Michael Maes, M.D., Ph.D., Sichuan Provincial Center for Mental Health, Sichuan Provincial People’s Hospital, School of Medicine, University of Electronic Science and Technology of China, Chengdu 610072, China. Co-Joint first authors (Min Wang, Mengqi Niu).

## Abstract

**Background:** Insomnia is highly prevalent in major depressive disorder (MDD), yet the clinical and biological significance of dysfunctional beliefs and attitudes about sleep (DBAS) in MDD remains poorly understood. We examined whether distinct DBAS dimensions are associated with insomnia severity, depressive phenotypes, and neuroimmune–metabolic–oxidative (NIMETOX) biomarkers.

**Methods:** DBAS-30 dimensions, clinical phenotypes, and NIMETOX biomarkers were assessed in 87 hospitalized patients with MDD and 31 healthy controls. Principal component analysis (PCA) was used to ascertain DBAS cognitive dimensions, and associations were examined using multivariable analyses.

**Results:** Compared with controls, MDD patients showed lower scores on “perceived consequences of insomnia,” “sleep-related worry and helplessness,” and “medication-related beliefs,” but higher “causal attributions to aging or biochemical imbalance” scores. A composite DBAS score reflecting these 4 MDD-associated cognitive dimensions was constructed, labeled DBAS dimensional index (DBAS-DI). This score was lower in MDD and inversely associated with Insomnia Severity Index (ISI) scores, overall MDD severity, physiosomatic symptoms, current suicidal ideation, and recurrence of illness. DBAS-DI and the expectations domain jointly explained 35.4% of the variance in ISI scores, with inverse and positive associations, respectively. The DBAS-DI score was inversely associated with indices of the acute-phase inflammatory response, oxidative stress, fecal branched-chain short-chain fatty acids (SCFAs) and positively associated with serum antioxidant capacity and protective SCFAs, such as acetate, propionate, and butyrate.

**Conclusions:** DBAS abnormalities in MDD extend beyond insomnia-related cognitions and are associated with illness severity, suicidality, recurrence, and NIMETOX alterations. These findings suggest that dysfunctional sleep-related cognitions are embedded within broader immune–oxidative and gut microbial–metabolic processes that may contribute to the insomnia–depression interface.

## Introduction

Major Depressive Disorder (MDD) affects approximately 236 million individuals worldwide and imposes a substantial global disease burden^1^. Prospective cohort data indicate that the lifetime prevalence of MDD exceeds 30% ^2^, with a 12-month prevalence ranging between 4% and 5% ^3^. Insomnia is a cardinal symptom during acute episodes and manifests in 80%–100% of patients with MDD^4,5^. Moreover, it frequently persists as a residual symptom following remission^6^, significantly elevates the risk of relapse^7,8^, thereby contributing to a chronic, fluctuating illness course. Furthermore, insomnia constitutes an independent risk factor for suicidal ideation (SI)^9,10^. Notably, residual symptoms, including persistent insomnia and mood disturbances, often endure among patients receiving effective antidepressant treatment or those who have achieved symptomatic remission^11^. Consequently, elucidating the underlying mechanisms that maintain these residual symptoms is imperative.

The Dysfunctional Beliefs and Attitudes about Sleep (DBAS) questionnaire, originally developed by Morin and colleagues^12,13^, provides a standardized instrument for assessing the core cognitive dimensions of insomnia. The original 30-item questionnaire encompasses several cognitive domains, including higher scores on causal attributions of insomnia to aging or biochemical imbalance^14^. Its brief version, the DBAS-16, distinguishes four domains: perceived consequences of insomnia, worry/helplessness, sleep expectations, and medication-related beliefs^15^. Carney and Edinger^14^ noted that good sleepers may also endorse dysfunctional sleep-related beliefs but generally do so less strongly. Stronger endorsement may reflect greater cognitive rigidity and vulnerability to sleep-related distress. Previous research has documented elevated DBAS-16 scores in patients with MDD compared with good sleepers^16^. Individuals with insomnia and depressive symptoms have also reported more dysfunctional sleep-related beliefs than those with insomnia without significant depressive symptoms^17^. Cognitive Behavioral Therapy for Insomnia (CBT-I) has been demonstrated to effectively ameliorate dysfunctional beliefs and attitudes about sleep (DBAS)^18,19^. Faaland et al.^20^ further reported that changes in DBAS mediated the effects of digital CBT-I on psychological distress and insomnia severity.

Accumulating evidence indicates^21,22^ that neuro-immune-metabolic-oxidative (NIMETOX) pathways are implicated in MDD and its progression. Activation of the peripheral immune-inflammatory response system (IRS) and consequent acute phase protein (APP) reprogramming, characterized by elevated positive APPs (e.g., monomeric C-reactive protein) and reduced negative APPs (e.g., albumin and transferrin)^23–25^, contribute to diminished antioxidant and anti-inflammatory capacity. This milieu drives oxidative and nitrosative stress, lipid peroxidation, and lipotoxic remodeling, leading to alterations in oxidative stress biomarkers. These alterations involve lipid oxidation markers, including oxidized high-density lipoprotein (oxHDL) cholesterol^21^. Additional abnormalities include altered stool short-chain fatty acid (SCFA) profiles, including increased branched-chain SCFAs (BSCFAs, isobutyric, isovaleric, and 2-methylbutyric acids) and reduced protective SCFAs (acetate, propionate, butyrate) in patients with MDD^21,26^. These peripheral abnormalities may contribute to the key domains of overall severity of depression (OSOD), physiosomatic symptoms, and recurrence of illness (ROI) through peripheral–central signaling pathways^27^, providing a rationale for examining their associations with dysfunctional sleep-related beliefs. Previous studies have shown objective impairments in antioxidant defenses and increased lipid peroxidation in insomnia superimposed upon a systemic inflammatory state inherent to MDD^28–30^. The microbiota-gut-brain axis also may play a role in insomnia^31^.

Within this pathological framework, the dysfunctional beliefs about sleep may not be merely cognitive epiphenomena. Rather, they may contribute to the persistence of sleep-related distress in MDD. On one hand, chronic oxidative stress, as well as other biological pathways in MDD (sympathetic hyperarousal, HPA axis overdrive, impaired neuroplasticity within prefrontal–limbic circuits) may erode emotional regulatory flexibility regarding sleep-related cognitions, potentially fostering the consolidation of catastrophic beliefs^28,32^. On the other hand, DBAS-driven pre-sleep anxiety and hyperarousal further prolong sleep latency, thereby potentially sustaining nocturnal increases of reactive oxygen species (ROS) and perpetuating inflammatory vulnerability^33^. Consequently, the persistence of dysfunctional sleep-related beliefs offers a novel, integrated perspective for understanding the mechanisms underlying MDD chronicity. However, the dimensional structure of DBAS in MDD, its associations with clinical phenotypes, and its relationships with gut-NIMETOX markers remain insufficiently characterized.

Accordingly, this study had three aims. First, we examined five DBAS dimensions using principal component analysis (PCA) and compared their scores by MDD status and insomnia severity. Second, we investigated associations between DBAS dimensions and clinical phenotypes, including insomnia severity, overall severity of depression (OSOD), physiosomatic symptoms, current suicidal ideation, and ROI. Third, we examined associations between DBAS dimensions and biological markers, including the acute-phase inflammatory index (API), antioxidant defenses, lipid oxidation indices, and fecal SCFAs.

## Methods

### Participants

This study enrolled 118 participants, comprising 87 hospitalized patients diagnosed with MDD and 31 healthy controls (HCs). The case-control cross-sectional research was conducted at the International NIMETOX Center, Mental Health Center of Sichuan Provincial People’s Hospital, Chengdu, China, between 2024 and 2025. Participants ranged in age from 18 to 65 years. HCs were recruited from hospital staff, their relatives, and friends, and matched to cases by age, sex, education, and body mass index (BMI). All MDD participants were recruited from psychiatric inpatients and were diagnosed with a current MDD according to DSM-5 depression criteria using the Structured Clinical Interview for DSM-5 and were included only if their Hamilton Depression Rating Scale (HAMD-21) score was greater than 18^34^. Written informed consent was obtained from all participants or their legally authorized representatives. The study protocol was approved by the Ethics Committee of Sichuan Provincial People’s Hospital (Ethical No. 2024-203).

The exclusion criteria for all participants included: (1) major psychiatric disorders (e.g., bipolar disorder, schizophrenia, autism spectrum disorders, psycho-organic disorders, eating disorders, substance use disorders excluding nicotine dependence, schizoaffective disorder); (2) recent infections (e.g., influenza-like illness, upper respiratory tract infection) within the past 3 months, or clinically significant allergic reactions (e.g., allergic rhinitis, urticaria, asthma exacerbation) within the past 1 month, or chronic infectious disease (e.g. HIV infection); (3) neurological disorders such as epilepsy, multiple sclerosis, stroke, and neurodegenerative diseases; (4) autoimmune or systemic inflammatory diseases (e.g., rheumatoid arthritis, inflammatory bowel disease, systemic lupus erythematosus, psoriasis), cancer, or type 1 diabetes; (5) current use of immunomodulatory drugs, including glucocorticosteroids; (6) recent use of analgesics; (7) personality disorders (antisocial or borderline) or developmental disorders; (8) recent surgery (within 3 months); (9) use of therapeutic doses of antioxidants or omega-3 supplements within the previous 3 months; and (10) pregnancy or lactation. Controls were excluded if they had a history of MDD, dysthymia, anxiety disorders, or a familial history of mood disorders, substance use disorders (except nicotine dependence), or suicide.

### Clinical Assessments

All <u>subjects</u> had a systematic clinical assessment performed by a trained qualified physician, encompassing demographic, medical, psychiatric, and familial histories. The intensity of symptoms was evaluated on the same day utilizing standardized rating scales. The 21-item HAMD and the Hamilton Anxiety Rating Scale (HAMA) were used to assess the severity of depression and anxiety, respectively^35,36^. Self-reported depressed symptoms were evaluated using the Beck Depression Inventory (BDI-II)^37^, while state anxiety was quantified with the State-Trait Anxiety Inventory (STAI-state)^38^. Suicidal ideation and attempts, both historical and present, were evaluated with the Columbia Suicide Severity Rating Scale (C-SSRS)^39^. Insomnia severity was assessed using the Insomnia Severity Index (ISI)^40^, and Sleep-related beliefs and attitudes were assessed using the 30-item Dysfunctional Beliefs and Attitudes about Sleep questionnaire (DBAS-30), with the specific item content provided in **ESF Table 1**^13,41^. Items were rated from 1 (strongly agree) to 5 (strongly disagree), with higher scores indicating weaker endorsement of the statements. The assessment and calculation procedures for OSOD, physiosomatic symptoms, current SI, and ROI followed those described by Niu et al^42^. ACEs were assessed using the validated Chinese version of the Childhood Trauma Questionnaire Short Form (CTQ-SF)^43–45^. Body mass index (BMI) was calculated as weight in kilograms divided by height in meters squared.

**Table 1.** Demographic characteristics, clinical phenotypes, and biomarkers in patients with major depressive disorder (MDD) and healthy controls (HC).

| Variables | HC (n =31) | MDD (n = 87) | F/$\chi^2$/FET | df | P |
| --- | --- | --- | --- | --- | --- |
| Age (years) | 38.81(14.40) | 36.00 (12.89 ) | 1.02 | 1/116 | 0.32 |
| Sex (female/male) | 21/10 | 62/25 | 0.14 | 1 | 0.71 |
| BMI (kg/m<sup>2</sup>) | 23.44 (3.64 ) | 22.33 (3.22 ) | 2.53 | 1/116 | 0.12 |
| Education (years) | 13.48 (4.72 ) | 13.57 (3.43 ) | 0.01 | 1/116 | 0.91 |
| Married (yes/no) | 13/18 | 48/39 | 1.60 | 1 | 0.21 |
| Residence (village/city) | 4/27 | 6/81 | FET | - | 0.29 |
| Current smoking (yes/ no) | 1/30 | 13/74 | FET | - | 0.11 |
| Current drinking (yes/ no) | 6/25 | 8/79 | 2.26 | 1 | 0.13 |
| Total ISI | 4.13 (4.26 ) | 17.92 (6.21 ) | 130.51 | 1/115 | <0.001 |
| Total ACEs | 36.32 (7.58 ) | 46.52 (13.56 ) | 15.71 | 1/116 | <0.001 |
| Physiosomatic symptoms | -1.27 (0.43 ) | 0.50 (0.70 ) | 172.33 | 1/115 | <0.001 |
| Current SI | -0.76 (0.30 ) | 0.25 (1.01 ) | 30.00 | 1/115 | <0.001 |
| ROI | -2.53 (0.38 ) | 0.92 (2.22 ) | 73.62 | 1/115 | <0.001 |
| OSOD | -1.44 (0.27) | 0.54 (0.53) | 390.68 | 1/115 | <0.001 |
| Total ANTIOX | 0.69 (0.63 ) | -0.27 (1.00 ) | 24.17 | 1/115 | <0.001 |
| z HDL – z OxHDL | 0.56 (0.56 ) | -0.15 (1.03 ) | 12.78 | 1/115 | 0.001 |
| z OxHDL – z OxLDL | -0.91 (0.80 ) | 0.13 (0.94 ) | 29.56 | 1/114 | <0.001 |
| API | -1.32 (1.97 ) | 0.45 (1.75 ) | 21.43 | 1/115 | <0.001 |
| Acetic acid | 0.56 (0.60 ) | -0.28 (1.10 ) | 15.25 | 1/95 | <0.001 |
| Propionic acid | 0.45 (0.62 ) | -0.29 (1.12 ) | 10.99 | 1/95 | 0.001 |
| Isobutyric acid | -0.10 (0.99 ) | -0.09 (1.11 ) | 0.002 | 1/95 | 0.96 |
| Butyric acid | 0.34 (0.69 ) | -0.11 (0.98 ) | 5.17 | 1/95 | 0.025 |
| Isovaleric acid | -0.41 (1.02 ) | 0.04 (1.08 ) | 3.72 | 1/95 | 0.057 |
| Pentanoic acid | 0.14 (0.94 ) | -0.20 (1.07 ) | 2.15 | 1/95 | 0.146 |
| Hexanoic acid | -0.19 (0.96 ) | -0.11 (0.96 ) | 0.13 | 1/95 | 0.72 |
| 2-Methylbutyric acid | -0.44 (1.03 ) | 0.01 (1.05 ) | 3.71 | 1/95 | 0.057 |
| Succinic acid | 0.29 (0.91 ) | -0.05 (1.10 ) | 2.22 | 1/95 | 0.14 |
| Protective SCFAs | 1.36 (1.64 ) | -0.68 (2.95 ) | 12.16 | 1/95 | 0.001 |
| BSCFA index | -0.75 (1.07 ) | 0.14 (1.04 ) | 14.60 | 1/95 | <0.001 |

Continuous variables are presented as mean (standard deviation), and categorical variables as counts. F, F statistic from analysis of variance; $\chi^2$ , Pearson's chi-square statistic; FET, Fisher's exact test; df, degrees of freedom (numerator/denominator for F tests). Sample sizes vary across variables because of missing data. BMI, body mass index; ISI, Insomnia Severity Index; ACEs, adverse childhood experiences; SI, suicidal ideation; ROI, recurrence of illness index; OSOD, overall severity of depression; Total ANTIOX, total antioxidant index; HDL, high-density lipoprotein cholesterol; OxHDL, oxidized high-density lipoprotein; OxLDL, oxidized low-density lipoprotein; API, acute-phase inflammatory index; SCFAs, short-chain fatty acids; BSCFA index, branched-chain fatty acid index; z, standardized score.

### Assays

Fasting venous blood was collected between 06:30 and 08:00, centrifuged at 3,500 rpm, and serum aliquots were stored at −80 °C until analysis. Fecal samples were also stored at −80 °C until analysis. Serum biomarkers were measured using biochemical, spectrophotometric, or enzyme-linked immunosorbent assays, whereas fecal short-chain fatty acids were quantified using UPLC–MS/MS. Detailed assay methods, kits, equipment, and analytical performance are provided in **ESF Table 2**. The components and calculation methods of the biomarker composite indices are presented in **ESF Table 3**.

**Table 2.** Principal components (PCs) analyses of the Dysfunctional Beliefs and Attitudes about Sleep (DBAS).

| PC_consequences |  | PC_worry |  | PC_expectations |  | PC_medications |  | PC_attributions |  |
| --- | --- | --- | --- | --- | --- | --- | --- | --- | --- |
| Variables | Loading | Variables | Loading | Variables | Loading | Variables | Loading | Variables | Loading |
| DBAS10 | 0.711 | DBAS5 | 0.693 | DBAS1 | 0.881 | DBAS11 | 0.837 | DBAS14 | 0.778 |
| DBAS12 | 0.712 | DBAS8 | 0.831 | DBAS2 | 0.881 | DBAS27 | 0.837 | DBAS24 | 0.778 |
| DBAS18 | 0.833 | DBAS17 | 0.701 |  |  |  |  |  |  |
| DBAS21 | 0.727 | DBAS19 | 0.650 |  |  |  |  |  |  |
| DBAS30 | 0.741 | DBAS20 | 0.690 |  |  |  |  |  |  |
|  |  | DBAS25 | 0.777 |  |  |  |  |  |  |
| KMO = 0.710<br>X<sup>2</sup> = 204.985<br>df=10, p<0.001<br>VE =55.649% |  | KMO = 0.798<br>X<sup>2</sup>=224.261<br>df=15, p<0.001<br>VE =52.771% |  | KMO = 0.5<br>X<sup>2</sup> =41.800<br>df=1, p<0.001<br>VE = 77.552% |  | KMO = 0.5<br>X<sup>2</sup>= 20.367<br>df=1, p<0.001<br>VE = 70.104% |  | KMO = 0.5<br>X<sup>2</sup> = 5.296<br>df=1, p=0.021<br>VE = 60.585% |  |

PCA was performed separately within each item set. PC, principal component; DBAS, Dysfunctional Beliefs and Attitudes about Sleep; KMO, Kaiser–Meyer–Olkin measure of sampling adequacy; VE, variance explained; $\chi^2$ , chi-square statistic for Bartlett’s test of sphericity; df, degrees of freedom.

**Table 3.** Features of Dysfunctional beliefs and attitudes about sleep in patients with major depressive disorder (MDD) and healthy controls (HC)

| Variables | HC (n =31) | MDD (n = 87) | F | df | P |
| --- | --- | --- | --- | --- | --- |
| PC_consequences | 0.495 (0.171) | −0.176 (0.102) | 11.328 | 1/113 | 0.001 |
| PC_worry | 0.762 (0.156) | −0.271 (0.093) | 32.288 | 1/113 | <0.001 |
| PC_expectations | −0.030 (0.181) | 0.011 (0.108) | 0.038 | 1/113 | 0.846 |
| PC_medications | 0.556 (0.166) | −0.198 (0.099) | 15.089 | 1/113 | <0.001 |
| PC_attributions | −0.362 (0.167) | 0.129 (0.100) | 6.337 | 1/113 | 0.013 |
| DBAS-DI | 0.854 (0.153) | −0.304 (0.091) | 42.045 | 1/113 | <0.001 |

Values are estimated marginal means (standard errors) from analyses of covariance adjusted for sex, age, and years of education. DBAS, Dysfunctional Beliefs and Attitudes about Sleep. F, F statistic from analysis of covariance. df, degrees of freedom. DBAS-DI is the z-standardized composite of PC\_consequences + PC\_worry + PC\_medications − PC\_attributions.

### Statistical Analysis

Analyses were performed using IBM SPSS Statistics 30 for Windows. Variable distributions were examined, and log10, square-root, or rank-based transformations or Winsorization were applied as appropriate. Continuous and categorical variables were compared using analysis of variance (ANOVA) and Pearson’s chi-square tests, respectively. Associations between continuous variables were assessed using Pearson’s correlations.

DBAS item groupings were informed by previously described cognitive domains and exploratory analyses^14,15^. Principal component analysis (PCA) was performed separately within five item sets representing consequences, worry/helplessness, expectations, medication-related beliefs, and causal attributions. The Kaiser–Meyer–Olkin (KMO) measure, Bartlett’s test of sphericity, component loadings, and explained variance were examined. Principal component scores were derived and used in subsequent analyses. General linear models examined differences in DBAS scores by MDD status and insomnia severity (ISI ≤7 versus >7), adjusting for age, sex, BMI, education years, smoking status, and medication use as appropriate. Multivariable linear regression analyses examined associations of DBAS scores, ACEs, and biomarkers with clinical phenotypes, including OSOD, physiosomatic symptoms, current SI, ROI, and ISI. Separate models examined associations of biomarkers and ACEs with the DBAS scores, with sex, age, BMI, and years of education included as covariates where appropriate. Variable selection combined manual regression, forward stepwise regression (entry p = 0.05; removal p = 0.07) with biological relevance within the NIMETOX framework to obtain parsimonious, biologically interpretable models. The selected variables were entered simultaneously into multivariable linear regression models using the forced-entry method to estimate their mutually adjusted associations with each outcome. Model robustness was assessed using 1,000 bootstrap resamples, which yielded consistent statistical significance patterns, along with collinearity diagnostics and residual analyses. Heteroscedasticity was assessed using White’s test and the modified Breusch–Pagan test, and multicollinearity using tolerance and variance inflation factors (VIFs). Standardized regression coefficients (β), degrees of freedom, p-values, R², and F statistics were reported. All tests were two-tailed, with statistical significance set at α = 0.05.

## Results

### Socio-demographic characteristics, clinical phenotypes, and biomarkers

**Table 1** summarizes the socio-demographic characteristics, clinical phenotypes, and biomarkers in patients with MDD and HCs. The groups did not differ significantly in age, sex, BMI, years of education, marital status, residence, current smoking, or alcohol use. Compared with HCs, patients with MDD had higher scores for ISI, Total ACEs, physiosomatic symptoms, current SI, ROI, and OSOD. Regarding biomarkers, patients with MDD had higher z OxHDL − z OxLDL, API, and BSCFA index scores, but lower Total ANTIOX, z HDL − z OxHDL, acetic acid, propionic acid, butyric acid, and protective SCFAs. Isobutyric acid, isovaleric acid, pentanoic acid, hexanoic acid, 2-methylbutyric acid, and succinic acid did not differ significantly between groups.

### Principal component analysis of five DBAS cognitive domains

DBAS-30 items were initially grouped according to previously described cognitive domains. The consequences, worry/helplessness, expectations, and medication-related item sets followed the corresponding DBAS-16 groupings described by Morin et al.^15^, whereas items 14 and 24 formed a causal-attribution item set consistent with the DBAS-30 domain described by Carney and Edinger^14^. PCA was performed separately within each item set using data from the present study. During these analyses, item 24 was excluded from the medication-related set but retained in the causal-attribution set, yielding five components: PC_consequences, reflecting perceived adverse consequences of insomnia; PC_worry, reflecting sleep-related worry and helplessness; PC_expectations, reflecting unrealistic expectations about sleep requirements; PC_medications, reflecting beliefs about the necessity and benefits of sleep medication; and PC_attributions, reflecting attributions of insomnia to aging or biochemical imbalance. Higher PC_consequences, PC_worry, and PC_medications scores indicate weaker endorsement of dysfunctional sleep-related beliefs, whereas lower scores indicate stronger endorsement. The interpretation of PC_attributions follows its constituent items.

These components explained 55.649%, 52.771%, 77.552%, 70.104%, and 60.585% of the variance within their respective item sets. Component loadings ranged from 0.650 to 0.881 (**Table 2**). Therefore, the PC scores extracted from the items in these 5 domains were employed in other statistical analyses.

### DBAS scores by MDD status and insomnia severity

Patients with MDD had lower PC_consequences, PC_worry, PC_medications, and DBAS-DI scores and higher PC_attributions scores than HCs. PC_expectations did not differ significantly between groups (**Table 3**). Consequently, we have computed a composite DBAS score reflecting these 4 MDD-associated cognitive alterations as PC_consequences + PC_worry + PC_medications - higher PC_attributions (z composite score), labeled DBAS dysregulation index (DBAS-DI)). Comparisons by insomnia severity showed the same pattern. Participants with ISI > 7 had lower PC_consequences, PC_worry, PC_medications, and DBAS-DI scores and higher PC_attributions scores than those with ISI ≤ 7, whereas PC_expectations did not differ significantly. Total ACEs, physiosomatic symptoms, current SI, ROI, and OSOD scores were also higher in participants with ISI > 7 (**Table 4**).

**Table 4.** Clinical and DBAS scores in subjects with ISI≤7 and ISI>7.

| Variables | ISI≤7 (n =33) | ISI$>$7 (n = 84) | F | df | P |
| --- | --- | --- | --- | --- | --- |
| Total ACEs | −0.432 (0.159) | 0.179 (0.099) | 10.630 | 1/112 | 0.001 |
| physiosomatic symptoms | −1.106 (0.124) | 0.434 (0.078) | 110.937 | 1/112 | <0.001 |
| Current SI | −0.542 (0.153) | 0.213 (0.096) | 17.568 | 1/112 | <0.001 |
| ROI | −0.795 (0.139) | 0.312 (0.087) | 45.331 | 1/112 | <0.001 |
| OSOD | −1.211 (0.110) | 0.476 (0.069) | 167.839 | 1/112 | <0.001 |
| PC_consequences | 0.576 (0.160) | −0.202 (0.100) | 16.916 | 1/112 | <0.001 |
| PC_worry | 0.856 (0.141) | −0.315 (0.089) | 49.140 | 1/112 | <0.001 |
| PC_expectations | −0.062 (0.175) | 0.035 (0.110) | 0.218 | 1/112 | 0.641 |
| PC_medications | 0.511 (0.161) | −0.182 (0.101) | 13.215 | 1/112 | <0.001 |
| PC_attributions | −0.274 (0.161) | 0.135 (0.101) | 4.655 | 1/112 | 0.033 |
| DBAS-DI | 0.870 (0.144) | −0.328 (0.090) | 49.443 | 1/112 | <0.001 |

Values are estimated marginal means (standard errors) from analyses of covariance adjusted for sex, age, and years of education. DBAS-DI is the z-standardized composite of PC\_consequences + PC\_worry + PC\_medications − PC\_attributions. F, F statistic from analysis of covariance; df, degrees of freedom; PC, principal component; ISI, Insomnia Severity Index; ACEs, adverse childhood experiences; SI, suicidal ideation; ROI, recurrence of illness index; OSOD, overall severity of depression; DBAS, Dysfunctional Beliefs and Attitudes about Sleep.

Among the 87 patients with MDD, 59 used antidepressants, 9 used mood stabilizers, 28 used antipsychotics, and 40 used benzodiazepines. None of these medication classes were significantly associated with any of the five DBAS components or DBAS-DI, even before FDR correction. Antidepressant use was associated with z HDL − z OxHDL (F = 4.141, df = 1/61, p = 0.046), pentanoic acid (F = 5.547, df = 1/61, p = 0.022), and 2-methylbutyric acid (F = 4.328, df = 1/61, p = 0.042). However, none of these associations remained significant after Benjamini–Hochberg FDR correction (all adjusted p = 0.220). Mood stabilizer, antipsychotic, and benzodiazepine use were not significantly associated with any measured biomarker, even before FDR correction.

### Correlations of DBAS with clinical phenotypes, ACEs, and biomarkers

In all subjects, DBAS-DI, PC_worry, and PC_medications were negatively correlated with all five clinical phenotypes, whereas PC_consequences was negatively correlated with OSOD, ISI, and physiosomatic symptoms. Total ACEs were negatively correlated with DBAS-DI and PC_worry and positively correlated with PC_attributions, which also correlated positively with ROI and OSOD. PC_expectations showed no significant correlations with clinical phenotypes or ACEs (**Table 5**).

**Table 5.**
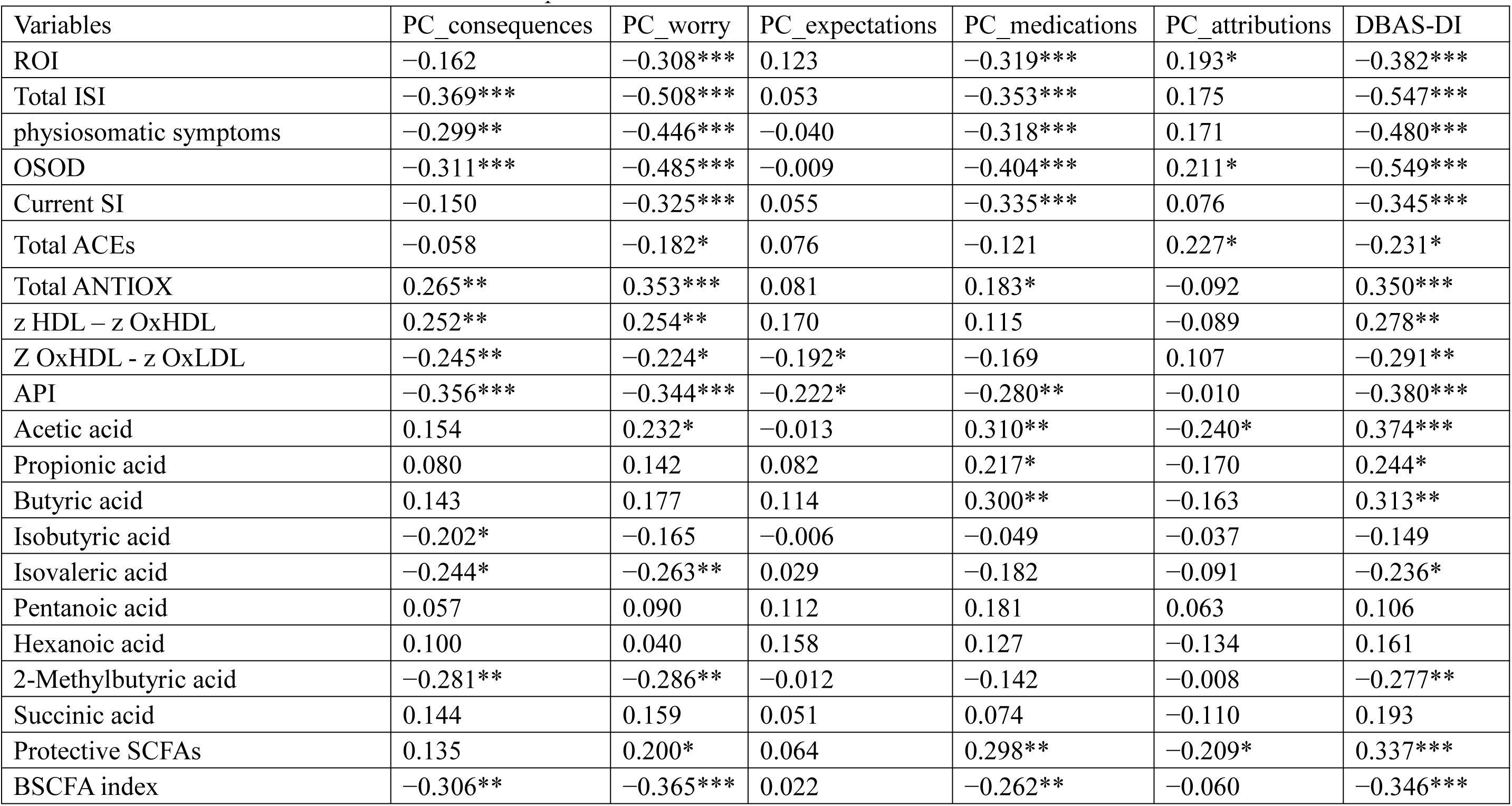
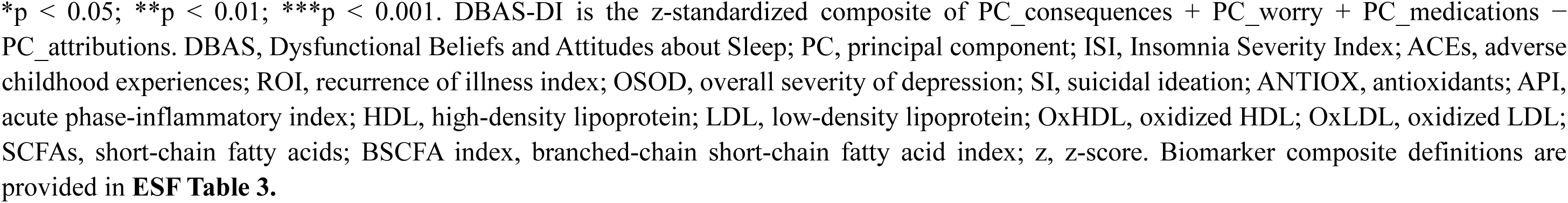
Correlations of DBAS scores with clinical phenome and biomarkers.

DBAS-DI was positively correlated with Total ANTIOX, z HDL − z OxHDL, acetic acid, propionic acid, butyric acid, and protective SCFAs, and negatively correlated with z OxHDL – z OxLDL, API, isovaleric acid, 2-methylbutyric acid, and the BSCFA index. PC_consequences, PC_worry, and PC_medications shared several of these associations. Biomarker correlations for PC_expectations were limited to negative correlations with z OxHDL − z OxLDL and API, and those for PC_attributions to negative correlations with acetic acid and protective SCFAs. No DBAS measure correlated significantly with pentanoic acid, hexanoic acid, or succinic acid.

### Multiple linear regression models of clinical phenotypes

**Table 6** summarizes the regression analyses with clinical phenotypes as dependent variables in all subjects. For OSOD, Model #1 explained 53.2% of the variance through Total ANTIOX (negative) and Total ACEs, z OxHDL − z OxLDL, and the BSCFA index (all positive). After adding DBAS measures as candidate variables, Model #2 additionally retained DBAS-DI (negative) and explained 55.6% of the variance.

**Table 6.**
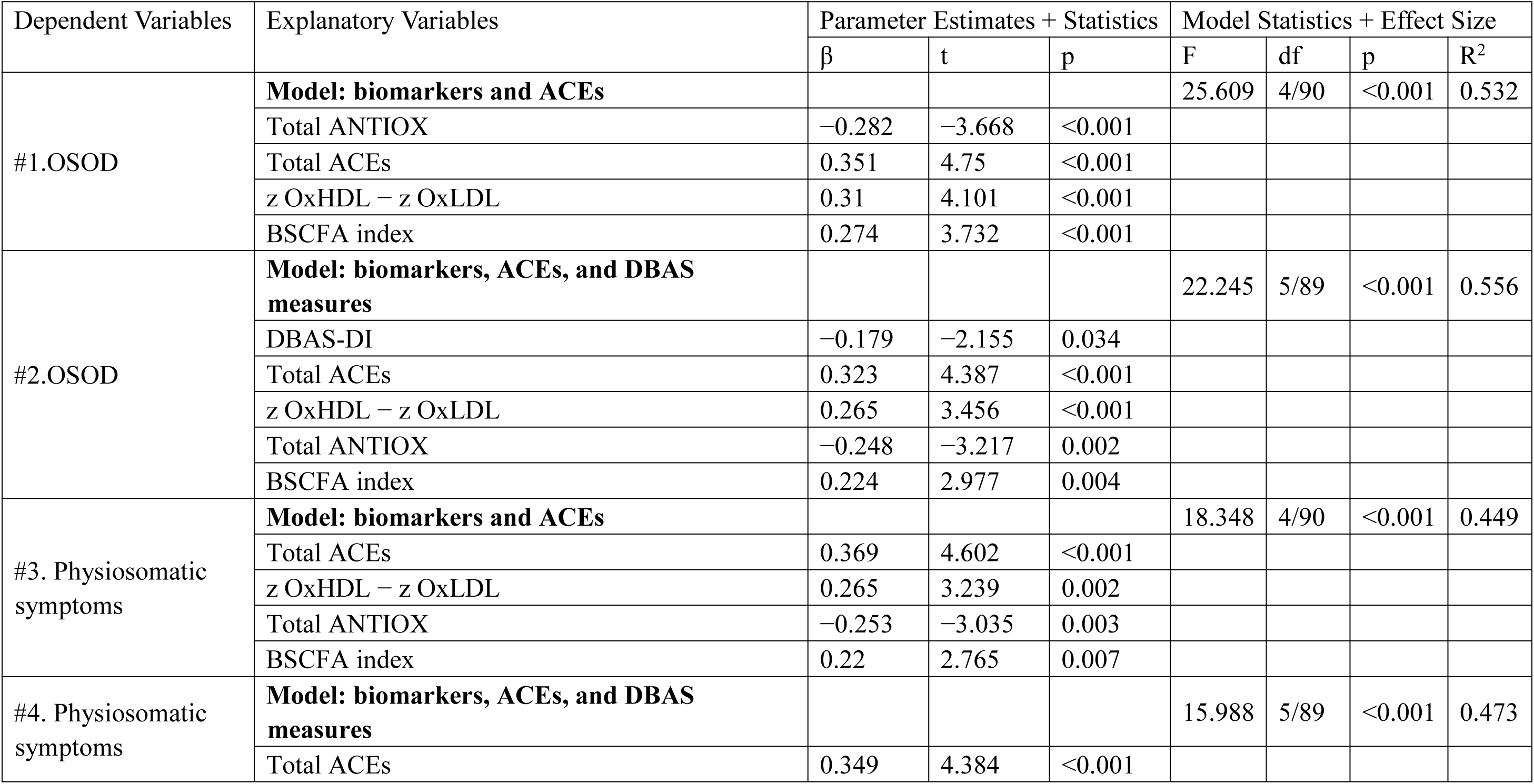
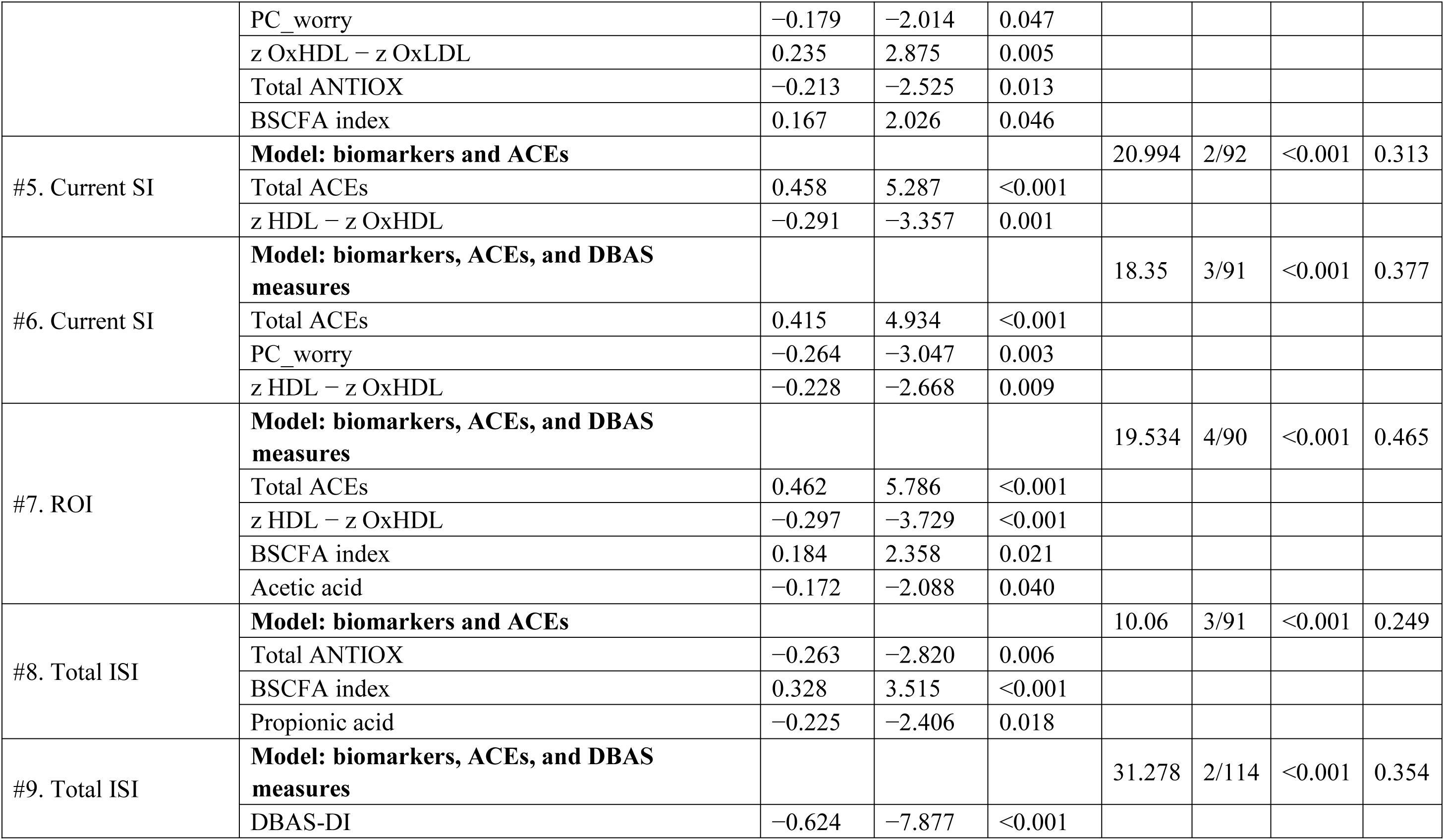
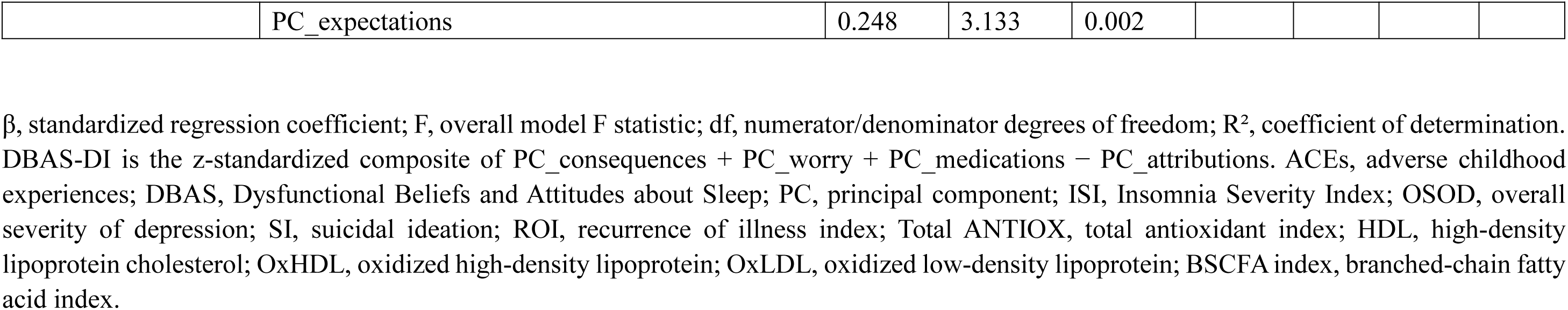
Multiple linear regression analyses of clinical phenome in relation to biomarkers, ACEs, and DBAS measures.

| Dependent Variables | Explanatory Variables | Parameter Estimates + Statistics |  |  | Model Statistics + Effect Size |  |  |  |
| --- | --- | --- | --- | --- | --- | --- | --- | --- |
| | | $\beta$ | t | p | F | df | p | R<sup>2</sup> |
| #1.OSOD | **Model: biomarkers and ACEs** |  |  |  | 25.609 | 4/90 | <0.001 | 0.532 |
|  | Total ANTIOX | -0.282 | -3.668 | <0.001 |  |  |  |  |
|  | Total ACEs | 0.351 | 4.75 | <0.001 |  |  |  |  |
|  | z OxHDL – z OxLDL | 0.31 | 4.101 | <0.001 |  |  |  |  |
|  | BSCFA index | 0.274 | 3.732 | <0.001 |  |  |  |  |
| #2.OSOD | **Model: biomarkers, ACEs, and DBAS measures** |  |  |  | 22.245 | 5/89 | <0.001 | 0.556 |
|  | DBAS-DI | -0.179 | -2.155 | 0.034 |  |  |  |  |
|  | Total ACEs | 0.323 | 4.387 | <0.001 |  |  |  |  |
|  | z OxHDL – z OxLDL | 0.265 | 3.456 | <0.001 |  |  |  |  |
|  | Total ANTIOX | -0.248 | -3.217 | 0.002 |  |  |  |  |
|  | BSCFA index | 0.224 | 2.977 | 0.004 |  |  |  |  |
| #3. Physiosomatic symptoms | **Model: biomarkers and ACEs** |  |  |  | 18.348 | 4/90 | <0.001 | 0.449 |
|  | Total ACEs | 0.369 | 4.602 | <0.001 |  |  |  |  |
|  | z OxHDL – z OxLDL | 0.265 | 3.239 | 0.002 |  |  |  |  |
|  | Total ANTIOX | -0.253 | -3.035 | 0.003 |  |  |  |  |
|  | BSCFA index | 0.22 | 2.765 | 0.007 |  |  |  |  |
| #4. Physiosomatic symptoms | **Model: biomarkers, ACEs, and DBAS measures** |  |  |  | 15.988 | 5/89 | <0.001 | 0.473 |
|  | Total ACEs | 0.349 | 4.384 | <0.001 |  |  |  |  |
|  | PC_worry | -0.179 | -2.014 | 0.047 |  |  |  |  |
|  | z OxHDL – z OxLDL | 0.235 | 2.875 | 0.005 |  |  |  |  |
|  | Total ANTIOX | -0.213 | -2.525 | 0.013 |  |  |  |  |
|  | BSCFA index | 0.167 | 2.026 | 0.046 |  |  |  |  |
| #5. Current SI | **Model: biomarkers and ACEs** |  |  |  | 20.994 | 2/92 | <0.001 | 0.313 |
|  | Total ACEs | 0.458 | 5.287 | <0.001 |  |  |  |  |
|  | z HDL – z OxHDL | -0.291 | -3.357 | 0.001 |  |  |  |  |
| #6. Current SI | **Model: biomarkers, ACEs, and DBAS measures** |  |  |  | 18.35 | 3/91 | <0.001 | 0.377 |
|  | Total ACEs | 0.415 | 4.934 | <0.001 |  |  |  |  |
|  | PC_worry | -0.264 | -3.047 | 0.003 |  |  |  |  |
|  | z HDL – z OxHDL | -0.228 | -2.668 | 0.009 |  |  |  |  |
| #7. ROI | **Model: biomarkers, ACEs, and DBAS measures** |  |  |  | 19.534 | 4/90 | <0.001 | 0.465 |
|  | Total ACEs | 0.462 | 5.786 | <0.001 |  |  |  |  |
|  | z HDL – z OxHDL | -0.297 | -3.729 | <0.001 |  |  |  |  |
|  | BSCFA index | 0.184 | 2.358 | 0.021 |  |  |  |  |
|  | Acetic acid | -0.172 | -2.088 | 0.040 |  |  |  |  |
| #8. Total ISI | **Model: biomarkers and ACEs** |  |  |  | 10.06 | 3/91 | <0.001 | 0.249 |
|  | Total ANTIOX | -0.263 | -2.820 | 0.006 |  |  |  |  |
|  | BSCFA index | 0.328 | 3.515 | <0.001 |  |  |  |  |
|  | Propionic acid | -0.225 | -2.406 | 0.018 |  |  |  |  |
| #9. Total ISI | **Model: biomarkers, ACEs, and DBAS measures** |  |  |  | 31.278 | 2/114 | <0.001 | 0.354 |
|  | DBAS-DI | -0.624 | -7.877 | <0.001 |  |  |  |  |
|  | PC_expectations | 0.248 | 3.133 | 0.002 |  |  |  |  |

$\beta$ , standardized regression coefficient; F, overall model F statistic; df, numerator/denominator degrees of freedom; $R^2$ , coefficient of determination. DBAS-DI is the z-standardized composite of PC\_consequences + PC\_worry + PC\_medications – PC\_attributions. ACEs, adverse childhood experiences; DBAS, Dysfunctional Beliefs and Attitudes about Sleep; PC, principal component; ISI, Insomnia Severity Index; OSOD, overall severity of depression; SI, suicidal ideation; ROI, recurrence of illness index; Total ANTIOX, total antioxidant index; HDL, high-density lipoprotein cholesterol; OxHDL, oxidized high-density lipoprotein; OxLDL, oxidized low-density lipoprotein; BSCFA index, branched-chain fatty acid index.

For physiosomatic symptoms, Model #3 explained 44.9% of the variance through Total ACEs, z OxHDL − z OxLDL, and the BSCFA index (all positive) and Total ANTIOX (negative). After adding DBAS measures as candidate variables, Model #4 additionally retained PC_worry (negative) and explained 47.3% of the variance.

For current SI, Model #5 explained 31.3% of the variance through Total ACEs (positive) and z HDL − z OxHDL (negative). After adding DBAS measures as candidate variables, Model #6 additionally retained PC_worry (negative) and explained 37.7% of the variance. For ROI, Model #7 explained 46.5% of the variance through Total ACEs and the BSCFA index (both positive) and z HDL − z OxHDL and acetic acid (both negative). The final model remained unchanged after adding DBAS measures as candidate variables.

For ISI, Model #8 explained 24.9% of the variance through Total ANTIOX and propionic acid (both negative) and the BSCFA index (positive). After adding DBAS measures as candidate variables, Model #9 retained DBAS-DI (negative) and PC_expectations (positive), jointly explaining 35.4% of the variance.

### Multiple linear regression models of DBAS scores

**Table 7** summarizes the regression analyses with DBAS component scores and DBAS-DI as dependent variables all subjects. In Model #1, API, and the BSCFA index (both negative) and hexanoic acid (positive) jointly explained 21.8% of the variance in PC_consequences. In Model #2, the BSCFA index (negative) and Total ANTIOX and hexanoic acid (both positive) jointly explained 27.2% of the variance in PC_worry. In Model #3, z OxHDL − z OxLDL was negatively associated with PC_expectations, explaining 7.9% of its variance. In Model #4, acetic acid and years of education (both positive) jointly explained 19.7% of the variance in PC_medications. In Model #5, years of education and Total ACEs (both positive) jointly explained 13.4% of the variance in PC_attributions. In Model #6, API, and the BSCFA index (both negative) and hexanoic acid and z HDL − z OxHDL (both positive) jointly explained 38.6% of the variance in DBAS-DI.

**Table 7.**
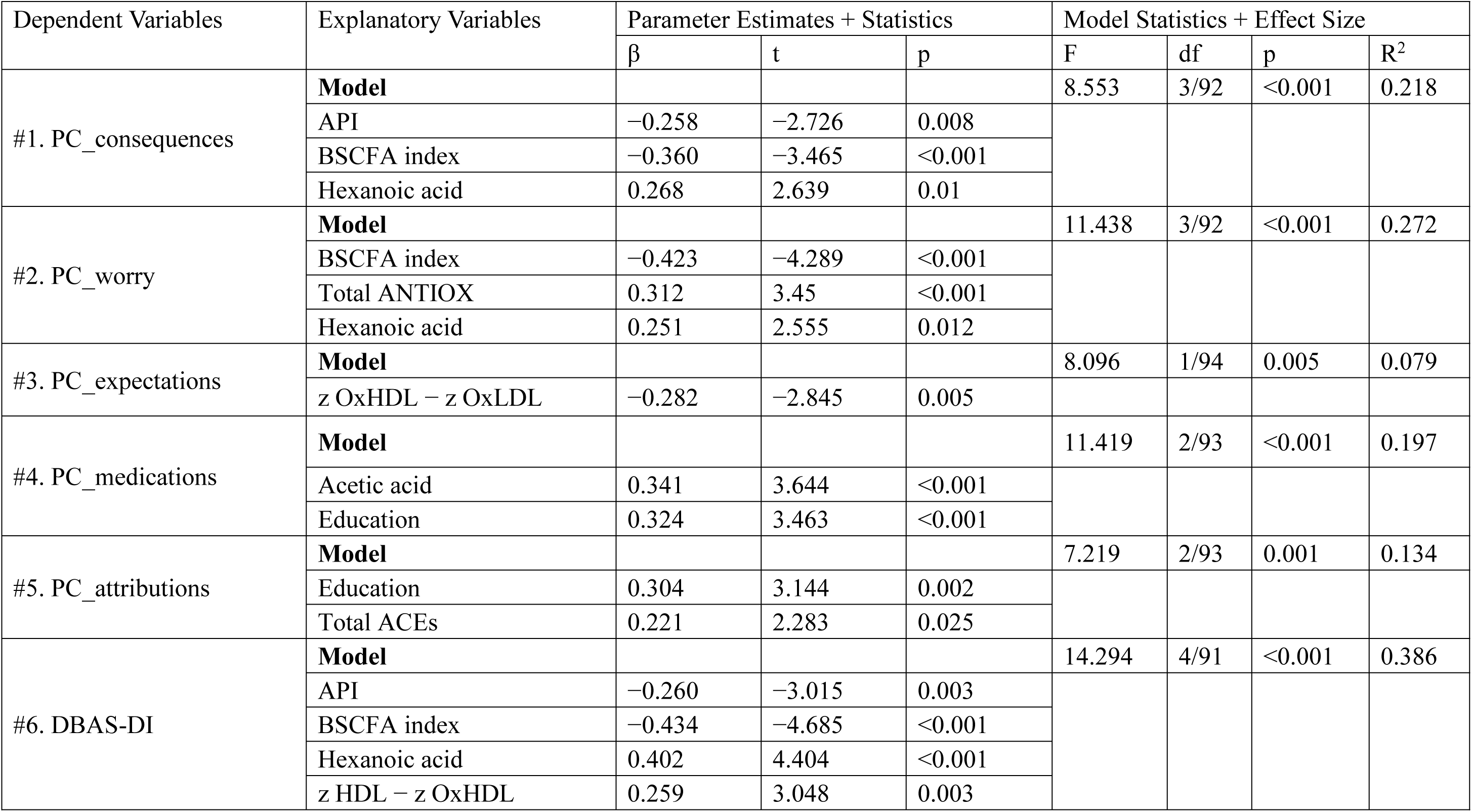
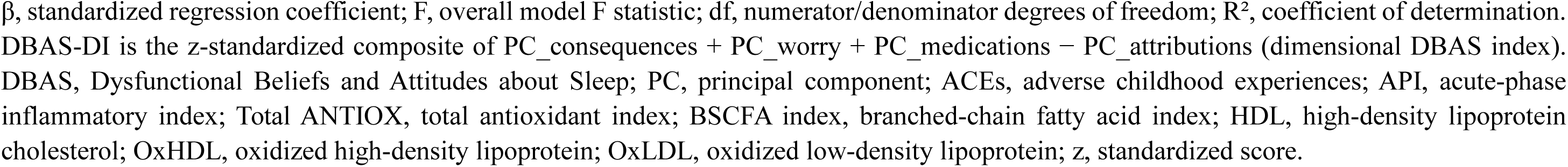
Multiple linear regression analyses of DBAS scores in relation to biomarkers and adverse childhood experiences.

## Discussion

### Five DBAS components in MDD

The first major finding of this study is that patients with MDD showed lower PC_consequences, PC_worry, and PC_medications scores and lower DBAS-DI scores than controls. In contrast, higher PC_attributions scores were observed in MDD patients. Higher DBAS aberration scores were associated with greater insomnia severity, overall MDD severity, physiosomatic symptoms, current suicidal ideation, and recurrence of illness. Moreover, DBAS-DI and PC_expectations jointly explained 35.4% of the variance in ISI scores, with DBAS-DI showing an inverse and PC_expectations a positive association with insomnia severity. Previous studies have reported elevated dysfunctional sleep-related beliefs in MDD, with higher DBAS-16 scores in patients with MDD than in good sleepers and in individuals with comorbid insomnia and depressive symptoms than in those with insomnia without significant depressive symptoms^16,17^.

Previous work supports the close relationship between dysfunctional sleep beliefs and subjective insomnia. Morin and colleagues^15^ reported that DBAS scores were more consistently related to self-reported insomnia severity, anxiety, and depressive symptoms than to objective polysomnographic measures, suggesting that the DBAS primarily captures the cognitive appraisal and subjective experience of disturbed sleep. Edinger and Wohlgemuth^46^ found that DBAS significantly differentiated patients with persistent insomnia from normal sleepers, with greater endorsement of dysfunctional sleep-related beliefs in the insomnia group.

The present study extends these observations to hospitalized patients with MDD: differences across DBAS dimensions between healthy controls and patients with MDD closely paralleled those observed between participants with lower and higher insomnia severity. These convergent patterns suggest that dysfunctional sleep-related beliefs are not merely characteristics of primary insomnia but are also prominent within the insomnia phenotype of MDD.

The clinical relevance of these associations is reinforced by previous treatment studies showing that dysfunctional sleep beliefs are not merely correlates of insomnia but may represent modifiable cognitive processes linked to treatment response^18–20,47^. Reductions in dysfunctional sleep beliefs have been implicated in improvements in insomnia severity, psychological distress, and fatigue following CBT-I^20,48^, and have been associated with improvements in both subjective and objective sleep outcomes and with the maintenance of these improvements during follow-up^48,49^. Nevertheless, previous studies have largely focused on DBAS total scores or broader subscales and have not delineated which specific DBAS cognitive dimensions are most strongly associated with the clinical phenotype of MDD.

The association of PC_worry with insomnia and depressive severity suggests that sleep-related worry may represent a particularly relevant cognitive dimension warranting further investigation. This is consistent with item-response analyses showing that worry- and helplessness-related items provide substantial information about the latent construct underlying dysfunctional beliefs and attitudes about sleep^50^. Persistent sleep-related worry could plausibly intensify this relationship through cognitive hyperarousal, perceived loss of control, fatigue, and anticipation of impaired daytime functioning. MDD and insomnia share physiological features of hyperarousal, including alterations in sympathetic and endocrine activity and increased fast-frequency EEG activity^51^. Increased beta and gamma activity during sleep has also been interpreted as potentially reflecting persistent worry, rumination, and arousal in MDD^52,53^. Thus, PC_worry could capture a cognitive manifestation of a broader hyperarousal phenotype rather than functioning as an isolated cognitive abnormality.

Moreover, greater depressive severity may amplify negative appraisal, hopelessness, rumination, and selective attention to inadequate sleep. Insomnia approximately doubles the subsequent risk of depression^54^ and frequently persists during MDD, supporting a bidirectional relationship in which sleep and depressive symptoms may mutually reinforce one another^55^. Poor sleep may further strengthen beliefs that sleep is uncontrollable^33,49^. Mood-congruent belief updating in depression^56^, whereby negative beliefs are insufficiently revised by positive information, may perpetuate this cycle^57,58^.

A similar process may underlie physiosomatic symptoms. Expectations of adverse physical consequences of poor sleep may increase attention to fatigue, pain, and low energy, thereby reinforcing dysfunctional sleep beliefs. In MDD, insomnia and physiosomatic symptoms may thus mutually reinforce one another through cognitive hyperarousal and rumination^55^, providing a plausible framework for the association with PC_worry.

The association between PC_worry and current suicidal ideation is particularly relevant. Sleep disturbances may predict subsequent suicidal thoughts and behaviors^59,60^, while worsening sleep may increase next-day suicidal ideation^61,62^. Hyperarousal may represent a shared feature of insomnia, MDD, and suicidal ideation^63^.

PC_expectations captures beliefs about required sleep duration and compensatory behaviors, such as needing eight hours of sleep or compensating through naps or earlier bedtimes and is conceptually distinct from worry about sleep control or daytime consequences. Previous studies found sleep expectations associated with diary-recorded time in bed but not insomnia severity^15^, whereas worry/helplessness showed a stronger relationship with insomnia severity^64^.

### Biological and psychosocial correlates of DBAS dimensions

The second major finding of our study is that DBAS dimensions showed distinct biological and psychosocial correlates. DBAS-DI, PC_consequences, and PC_worry were predominantly associated with the acute-phase inflammatory response, antioxidant capacity, lipid-oxidation indices, and SCFAs. These findings align with evidence that MDD involves interconnected immune-inflammatory, oxidative, metabolic, and lipid-regulatory abnormalities^21^, including HDL-associated antioxidant protection, lipid peroxidation^65^, and reduced antioxidant defenses^66,67^. SCFAs may provide an additional gut–brain interface through immune and metabolic signaling^26^. Given that sleep disturbances in MDD are likewise associated with inflammation, oxidative stress, metabolic dysfunction, and microbiota–gut–brain alterations^55^, our findings suggest that dysfunctional sleep cognitions are embedded within this broader biological dysregulation and may interact with MDD severity and disturbed sleep.

Higher PC_attributions scores were associated with greater ACE exposure and more years of education. Childhood adversity is an established risk factor for depression and subsequent sleep disturbances^68^ and has been associated with inflammatory alterations in MDD^69,70^. Sleep-related attributions may therefore partly reflect enduring cognitive schemas shaped by experiences of threat, unpredictability, and reduced perceived control. This interpretation is consistent with models of depressive belief formation, in which earlier experiences contribute to core beliefs that subsequently bias the processing of threatening, aversive, or uncertain information^56^. Education may further shape knowledge and expectations regarding sleep, medication, and physical health. Thus, causal attributions about insomnia appear to have psychosocial correlates distinct from those underlying sleep-related worry and perceived consequences.

The biological correlates of DBAS further inform its association with insomnia severity. Previous studies reported reduced antioxidant enzyme activities and total antioxidant status in insomnia^71^, as well as lower fecal SCFAs inversely associated with insomnia severity^72^. Although the final ISI model included only DBAS-DI and PC_expectations, these cognitive measures were themselves associated with biological indices. Notably, the BSCFA index was positively associated with insomnia severity but negatively with DBAS-DI. Thus, dysfunctional sleep cognitions and biological dysregulation may represent interrelated features of insomnia.

## Limitations

Several limitations should be considered when interpreting these findings. First, although the simultaneous assessment of suicidal ideation, recurrence indices, ACEs, multiple DBAS dimensions, and biological markers allowed sleep-related cognitions to be examined within an integrated clinical and biological framework, all patients with MDD were hospitalized during an acute depressive episode. The findings may therefore not generalize to community-based populations, patients with milder depression, or individuals in remission. Second, despite careful exclusion of major inflammatory and medical conditions, unmeasured lifestyle factors, including dietary intake and physical activity, may have influenced oxidative, lipid-related, and SCFA measures. Third, the observed associations between DBAS dimensions and antioxidants, lipid-oxidation, acute-phase protein, and SCFA-related indices should be considered preliminary. The single-center design and sample size warrant replication in independent cohorts in other cultures and countries. Finally, given the cross-sectional design, the temporal and causal relationships among dysfunctional sleep cognitions, insomnia severity, depressive symptoms, and biological alterations cannot be determined. Longitudinal and interventional studies are needed to establish whether these cognitive and biological abnormalities precede, follow, or reciprocally reinforce one another.

## Conclusions

In conclusion, dysfunctional beliefs and attitudes about sleep were prominent in MDD and were associated with insomnia severity and clinically important depressive phenotypes. Sleep-related worry was associated with physiosomatic symptoms and current suicidal ideation in multivariable analyses, whereas DBAS-DI was associated with OSOD and, together with PC_expectations, with insomnia severity. Moreover, DBAS components showed distinct associations with NIMETOX-related markers involving antioxidant defenses, acute-phase responses, lipid oxidation, and SCFA metabolism, and may represent clinically meaningful cognitive features embedded within broader interactions among insomnia, depressive psychopathology, and biological dysregulation. Future longitudinal and intervention studies should determine whether specifically targeting sleep-related cognitions, particularly sleep-related worry, can improve clinical outcomes and whether such improvement is accompanied by changes in relevant biological pathways.

## Data Availability

The database created during this investigation will be provided by the corresponding author (MM) upon a reasonable request once the authors have thoroughly used the dataset.

## Ethics approval

This study was approved by the Ethics Committee of Sichuan Provincial People’s Hospital (Ethics (Research) 2024-203), in compliance with the International Guidelines for Human Research Protection and the ethical principles outlined in the Declaration of Helsinki.

## Consent to participate

Before participating in this study, each subject provided written informed consent.

## Consent for publication

All authors have given their approval for this paper to be published.

## Author’s contributions

Min Wang: data curation, formal analysis, writing - original draft. Mengqi Niu: data curation, formal analysis, visualization, writing - original draft. Xinling Luo, Xiaoli Chen, Hongling Liu, Tangcong Chen: data curation. Abbas F. Almulla, Yingqian Zhang: writing - review and editing. Jia Li: data curation, writing - original draft. Michael Maes: supervision, conceptualization, formal analysis, writing - review and editing.

## References

1 Collaborators., G. M. D. Updated trends in the global prevalence and burden of mental disorders, 1990-2023: a systematic analysis for the Global Burden of Disease Study 2023. Lancet 407, 2040–2064, doi:10.1016/s0140-6736(26)00519-2 (2026).

2 Moffitt, T. E. et al. How common are common mental disorders? Evidence that lifetime prevalence rates are doubled by prospective versus retrospective ascertainment. Psychol Med 40, 899–909, doi:10.1017/s0033291709991036 (2010).

3 Marx, W. et al. Major depressive disorder. Nat Rev Dis Primers 9, 44, doi:10.1038/s41572-023-00454-1 (2023).

4 Dolsen, E. A., Asarnow, L. D. & Harvey, A. G. Insomnia as a transdiagnostic process in psychiatric disorders. Curr Psychiatry Rep 16, 471, doi:10.1007/s11920-014-0471-y (2014).

5 Geoffroy, P. A. et al. Insomnia and hypersomnia in major depressive episode: Prevalence, sociodemographic characteristics and psychiatric comorbidity in a population-based study. J Affect Disord 226, 132–141, doi:10.1016/j.jad.2017.09.032 (2018).

6 Carney, C. E., Segal, Z. V., Edinger, J. D. & Krystal, A. D. A comparison of rates of residual insomnia symptoms following pharmacotherapy or cognitive-behavioral therapy for major depressive disorder. J Clin Psychiatry 68, 254–260, doi:10.4088/jcp.v68n0211 (2007).

7 Dombrovski, A. Y. et al. Residual symptoms and recurrence during maintenance treatment of late-life depression. J Affect Disord 103, 77–82, doi:10.1016/j.jad.2007.01.020 (2007).

8 Ohayon, M. M. & Roth, T. Place of chronic insomnia in the course of depressive and anxiety disorders. J Psychiatr Res 37, 9–15, doi:10.1016/s0022-3956(02)00052-3 (2003).

9 Pigeon, W. R., Pinquart, M. & Conner, K. Meta-analysis of sleep disturbance and suicidal thoughts and behaviors. J Clin Psychiatry 73, e1160–1167, doi:10.4088/JCP.11r07586 (2012).

10 Johnson, E. O., Roth, T. & Breslau, N. The association of insomnia with anxiety disorders and depression: exploration of the direction of risk. J Psychiatr Res 40, 700–708, doi:10.1016/j.jpsychires.2006.07.008 (2006).

11 Zajecka, J., Kornstein, S. G. & Blier, P. Residual symptoms in major depressive disorder: prevalence, effects, and management. J Clin Psychiatry 74, 407–414, doi:10.4088/JCP.12059ah1 (2013).

12 Morin C, M. Insomnia: Psychological Assessment and Management. (Guilford Press, 1993).

13 Morin, C. M., Stone, J., Trinkle, D., Mercer, J. & Remsberg, S. Dysfunctional beliefs and attitudes about sleep among older adults with and without insomnia complaints. Psychol Aging 8, 463–467, doi:10.1037//0882-7974.8.3.463 (1993).

14 Carney, C. E. & Edinger, J. D. Identifying critical beliefs about sleep in primary insomnia. Sleep 29, 342–350, doi:10.1093/sleep/29.3.342 (2006).

15 Morin, C. M., Vallières, A. & Ivers, H. Dysfunctional beliefs and attitudes about sleep (DBAS): validation of a brief version (DBAS-16). Sleep 30, 1547–1554, doi:10.1093/sleep/30.11.1547 (2007).

16 Carney, C. E., Edinger, J. D., Manber, R., Garson, C. & Segal, Z. V. Beliefs about sleep in disorders characterized by sleep and mood disturbance. J Psychosom Res 62, 179–188, doi:10.1016/j.jpsychores.2006.08.006 (2007).

17 Levenson, J. C., Benca, R. M. & Rumble, M. E. Sleep Related Cognitions in Individuals with Symptoms of Insomnia and Depression. J Clin Sleep Med 11, 847–854, doi:10.5664/jcsm.4930 (2015).

18 Thakral, M., Von Korff, M., McCurry, S. M., Morin, C. M. & Vitiello, M. V. Changes in dysfunctional beliefs about sleep after cognitive behavioral therapy for insomnia: A systematic literature review and meta-analysis. Sleep Med Rev 49, 101230, doi:10.1016/j.smrv.2019.101230 (2020).

19 Song, Y. et al. Change in Dysfunctional Sleep-Related Beliefs is Associated with Changes in Sleep and Other Health Outcomes Among Older Veterans With Insomnia: Findings From a Randomized Controlled Trial. Ann Behav Med 56, 35–49, doi:10.1093/abm/kaab030 (2022).

20 Faaland, P. et al. Dysfunctional beliefs and attitudes about sleep (DBAS) mediate outcomes in dCBT-I on psychological distress, fatigue, and insomnia severity. Sleep Med 110, 1–6, doi:10.1016/j.sleep.2023.07.018 (2023).

21 Maes, M., Almulla, A. F., You, Z. & Zhang, Y. Neuroimmune, metabolic and oxidative stress pathways in major depressive disorder. Nat Rev Neurol 21, 473–489, doi:10.1038/s41582-025-01116-4 (2025).

22 Maes, M. et al. Key factors underpinning neuroimmune-metabolic-oxidative (NIMETOX) major depression in outpatients: paraoxonase 1 activity, reverse cholesterol transport, increased atherogenicity, protein oxidation, and differently expressed cytokine networks. Neuro Endocrinol Lett 46, 115–125 (2025).

23 Maes, M. et al. Anthropometric and biochemical assessment of the nutritional state in depression: evidence for lower visceral protein plasma levels in depression. J Affect Disord 23, 25–33, doi:10.1016/0165-0327(91)90032-n (1991).

24 Maes, M. A review on the acute phase response in major depression. Rev Neurosci 4, 407–416, doi:10.1515/revneuro.1993.4.4.407 (1993).

25 Almulla, A. F., Niu, M., Stoyanov, D., Zhang, Y. & Maes, M. Monomeric CRP and negative acute phase proteins, but not pentameric CRP, as biomarkers of major depression and MDMD. Acta Neuropsychiatr 38, e4, doi:10.1017/neu.2025.10044 (2025).

26 Niu, M. et al. A shift toward proteolytic gut fermentation links systemic inflammation to clinical phenotypes in major depressive disorder. Prog Neuropsychopharmacol Biol Psychiatry 149, 111900, doi:10.1016/j.pnpbp.2026.111900 (2026).

27 Estrada-Valbuena., et al. The Emerging Role of Microbial Extracellular Vesicles in Neuroinflammation and Neurodegeneration. Medicine Bulletin 2, 231–238, 10.1002/mdb2.70031 (2026).

28 Gulec, M. et al. Oxidative stress in patients with primary insomnia. Prog Neuropsychopharmacol Biol Psychiatry 37, 247–251, doi:10.1016/j.pnpbp.2012.02.011 (2012).

29 Yin, J. et al. Associations between sleep disturbance, inflammatory markers and depressive symptoms: Mediation analyses in a large NHANES community sample. Prog Neuropsychopharmacol Biol Psychiatry 126, 110786, doi:10.1016/j.pnpbp.2023.110786 (2023).

30 Fang, H., Tu, S., Sheng, J. & Shao, A. Depression in sleep disturbance: A review on a bidirectional relationship, mechanisms and treatment. J Cell Mol Med 23, 2324–2332, doi:10.1111/jcmm.14170 (2019).

31 Yang, M., Chen, Q., Meng, Z., Gu, X. & Bai, C. The Microbiota-Gut-Brain Axis in Insomnia: Mechanisms and Intervention Strategies. Life (Basel) 16, doi:10.3390/life16040583 (2026).

32 Irwin, M. R. Insomnia and Inflammation Conspire to Heighten Depression Risk: Implications for Treatment and Prevention of Mood Disorders. Biol Psychiatry 98, 819–829, doi:10.1016/j.biopsych.2025.04.018 (2025).

33 Harvey, A. G. A cognitive model of insomnia. Behav Res Ther 40, 869–893, doi:10.1016/s0005-7967(01)00061-4 (2002).

34 Maes, M. et al. Peripheral metabolic-redox signaling as a core mechanism of major depressive disorder: evidence from deep metabolomic phenotyping. Acta Neuropsychiatr 38, e26, doi:10.1017/neu.2026.10065 (2026).

35 Hamilton, M. The assessment of anxiety states by rating. Br J Med Psychol 32, 50–55, doi:10.1111/j.2044-8341.1959.tb00467.x (1959).

36 Hamilton, M. A rating scale for depression. J Neurol Neurosurg Psychiatry 23, 56–62, doi:10.1136/jnnp.23.1.56 (1960).

37 Beck, A. T., Steer, R. A., Ball, R. & Ranieri, W. Comparison of Beck Depression Inventories -IA and -II in psychiatric outpatients. J Pers Assess 67, 588–597, doi:10.1207/s15327752jpa6703_13 (1996).

38 Spielberger, C. D., Gorsuch, R. L., Lushene, R., Vagg, P. R. & Jacobs, G. A. Manual for the State-Trait Anxiety Inventory. (Consulting Psychologists Press., 1983).

39 Posner, K. et al. The Columbia-Suicide Severity Rating Scale: initial validity and internal consistency findings from three multisite studies with adolescents and adults. Am J Psychiatry 168, 1266–1277, doi:10.1176/appi.ajp.2011.10111704 (2011).

40 Bastien, C. H., Vallières, A. & Morin, C. M. Validation of the Insomnia Severity Index as an outcome measure for insomnia research. Sleep Med 2, 297–307, doi:10.1016/s1389-9457(00)00065-4 (2001).

41 Xu, B., Yang, L. & Li, Z. Reliability and validity of the Chinese Version Dysfunctional Beliefs and Attitudes about sleep scale. Journal of Capital Medical University 40, 665–670, doi:10.3969/j.issn.1006-7795.2019.05.001. (2019).

42 Niu, M. et al. Dissection of the clinical phenome of major depressive disorder into subdomains and subgroups in relation to activated immune-inflammatory profiles. medRxiv, 2025.2010.2021.25338439, doi:10.1101/2025.10.21.25338439 (2025).

43 Bernstein, D. P. et al. Development and validation of a brief screening version of the Childhood Trauma Questionnaire. Child Abuse Negl 27, 169–190, doi:10.1016/s0145-2134(02)00541-0 (2003).

44 Vasupanrajit, A., Maes, M., Jirakran, K. & Tunvirachaisakul, C. Complex Intersections Between Adverse Childhood Experiences and Negative Life Events Impact the Phenome of Major Depression. Psychology research and behavior management 17, 2161–2178, doi:10.2147/prbm.S458257 (2024).

45 Zhao, X. et al. Reliability and validity of the Chinese version of childhood trauma questionnaire. Chinese Journal of Clinical Rehabilitation 9, 105–107 (2005).

46 Edinger, J. D. & Wohlgemuth, W. K. Psychometric comparisons of the standard and abbreviated DBAS-10 versions of the dysfunctional beliefs and attitudes about sleep questionnaire. Sleep Med 2, 493–500, doi:10.1016/s1389-9457(01)00078-8 (2001).

47 Parsons, C. E., Zachariae, R., Landberger, C. & Young, K. S. How does cognitive behavioural therapy for insomnia work? A systematic review and meta-analysis of mediators of change. Clin Psychol Rev 86, 102027, doi:10.1016/j.cpr.2021.102027 (2021).

48 Edinger, J. D., Wohlgemuth, W. K., Radtke, R. A., Marsh, G. R. & Quillian, R. E. Does cognitive-behavioral insomnia therapy alter dysfunctional beliefs about sleep? Sleep 24, 591–599, doi:10.1093/sleep/24.5.591 (2001).

49 Morin, C. M., Blais, F. & Savard, J. Are changes in beliefs and attitudes about sleep related to sleep improvements in the treatment of insomnia? Behav Res Ther 40, 741–752, doi:10.1016/s0005-7967(01)00055-9 (2002).

50 Castillo, L. I. R., Hadjistavropoulos, T., Tan, L. O. & MacNab, Y. C. Item response theory analysis of the Dysfunctional Beliefs and Attitudes about Sleep-16 (DBAS-16) scale in a university student sample. PLoS One 18, e0281364, doi:10.1371/journal.pone.0281364 (2023).

51 Riemann, D., Krone, L. B., Wulff, K. & Nissen, C. Sleep, insomnia, and depression. Neuropsychopharmacology 45, 74–89, doi:10.1038/s41386-019-0411-y (2020).

52 Tekell, J. L. et al. High frequency EEG activity during sleep: characteristics in schizophrenia and depression. Clin EEG Neurosci 36, 25–35, doi:10.1177/155005940503600107 (2005).

53 Armitage, R. Sleep and circadian rhythms in mood disorders. Acta Psychiatr Scand Suppl, 104–115, doi:10.1111/j.1600-0447.2007.00968.x (2007).

54 Baglioni, C. et al. Insomnia as a predictor of depression: a meta-analytic evaluation of longitudinal epidemiological studies. J Affect Disord 135, 10–19, doi:10.1016/j.jad.2011.01.011 (2011).

55 Rosenblum, Y. et al. Sleep Neurophysiology in Depression. Biol Psychiatry 98, 842–853, doi:10.1016/j.biopsych.2025.07.023 (2025).

56 Bottemanne, H. et al. How mood shapes belief updating bias in depression. Cogn Affect Behav Neurosci 25, 889–903, doi:10.3758/s13415-025-01297-x (2025).

57 Korn, C. W., Sharot, T., Walter, H., Heekeren, H. R. & Dolan, R. J. Depression is related to an absence of optimistically biased belief updating about future life events. Psychol Med 44, 579–592, doi:10.1017/s0033291713001074 (2014).

58 Kube, T., Kirchner, L., Gärtner, T. & Glombiewski, J. A. How negative mood hinders belief updating in depression: results from two experimental studies. Psychol Med 53, 1288–1301, doi:10.1017/s0033291721002798 (2023).

59 Liu, R. T. et al. Sleep and suicide: A systematic review and meta-analysis of longitudinal studies. Clin Psychol Rev 81, 101895, doi:10.1016/j.cpr.2020.101895 (2020).

60 Harris, L. M., Huang, X., Linthicum, K. P., Bryen, C. P. & Ribeiro, J. D. Sleep disturbances as risk factors for suicidal thoughts and behaviours: a meta-analysis of longitudinal studies. Sci Rep 10, 13888, doi:10.1038/s41598-020-70866-6 (2020).

61 Wang, X., Cheng, S. & Xu, H. Systematic review and meta-analysis of the relationship between sleep disorders and suicidal behaviour in patients with depression. BMC Psychiatry 19, 303, doi:10.1186/s12888-019-2302-5 (2019).

62 Bourke, M., Sippel, F. & Fortnum, K. Sleep disturbances as a short-term within-person risk-factor for suicidal ideation: A systematic review and meta-analysis. J Psychiatr Res 199, 104–112, doi:10.1016/j.jpsychires.2026.04.033 (2026).

63 Dolsen, E. A. et al. Neurophysiological correlates of suicidal ideation in major depressive disorder: Hyperarousal during sleep. J Affect Disord 212, 160–166, doi:10.1016/j.jad.2017.01.025 (2017).

64 Cha, E. J., Hong, S., Kim, S., Chung, S. & Jeon, H. J. Contribution of dysfunctional sleep-related cognitions on insomnia severity: a network perspective. J Clin Sleep Med 20, 743–751, doi:10.5664/jcsm.11006 (2024).

65 Almulla, A. F. et al. Reverse cholesterol transport and lipid peroxidation biomarkers in major depression and bipolar disorder: A systematic review and meta-analysis. Brain, behavior, and immunity 113, 374–388, doi:10.1016/j.bbi.2023.08.007 (2023).

66 Jiménez-Fernández, S. et al. Oxidative stress and antioxidant parameters in patients with major depressive disorder compared to healthy controls before and after antidepressant treatment: results from a meta-analysis. J Clin Psychiatry 76, 1658–1667, doi:10.4088/JCP.14r09179 (2015).

67 Liu, T. et al. A Meta-Analysis of Oxidative Stress Markers in Depression. PLoS One 10, e0138904, doi:10.1371/journal.pone.0138904 (2015).

68 Yu, H. J., Liu, X., Yang, H. G., Chen, R. & He, Q. Q. The association of adverse childhood experiences and its subtypes with adulthood sleep problems: A systematic review and meta-analysis of cohort studies. Sleep Med 98, 26–33, doi:10.1016/j.sleep.2022.06.006 (2022).

69 Gill, H. et al. The Association Between Adverse Childhood Experiences and Inflammation in Patients with Major Depressive Disorder: A Systematic Review. J Affect Disord 272, 1–7, doi:10.1016/j.jad.2020.03.145 (2020).

70 Zagaria, A. et al. Inflammation as a mediator between adverse childhood experiences and adult depression: A meta-analytic structural equation model. J Affect Disord 357, 85–96, doi:10.1016/j.jad.2024.04.072 (2024).

71 Liang, B., Li, Y. H. & Kong, H. Serum paraoxonase, arylesterase activities and oxidative status in patients with insomnia. Eur Rev Med Pharmacol Sci 17, 2517–2522 (2013).

72 Yerlikaya, F. H. et al. Insomnia patients have a poor intestinal prognosis: Accompanied by microbiota-derived short chain fatty acids, diet and zonulin. J Psychiatr Res 183, 25–30, doi:10.1016/j.jpsychires.2025.01.035 (2025).

